# Temporal Network Dynamics and Boolean Modeling Reveal Critical State Transitions and Regulatory Hubs in Preeclampsia Pathogenesis

**DOI:** 10.1101/2025.08.08.25333339

**Authors:** Leandro Henrique Manfredi

## Abstract

**Background:** Preeclampsia (PE) is a severe hypertensive disorder of pregnancy driven by placental dysfunction. A complex interplay of inflammatory, metabolic, and endothelial pathways regulates the transition from a healthy pregnancy to a pathological state. However, the systems-level dynamics and the mechanisms that the predisposing factors determine disease’s onset and progression remain to be uncovered.

**Methodology:** An integrative systems biology approach was used to develop a representative PE time-aware network, using a temporal transcriptomic dataset from uncomplicated gestation. By mapping a curated PPI of PE onto normal placental DEGs, a dynamic, 30-node Boolean network model was constructed with rules curated from established signaling pathways. The main outcome that was used to represent a PE system state was the inactivation of *eNOS*. The model was used to identify stable cellular states (attractors) and to simulate the response to systemic conditions known to be involved in pathology, including inflammation, hypoxia, oxidative stress and metabolic overload (oxLDL). Complementary network diffusion analysis was performed expanding the PE-time-aware direct network to identify key propagating pathways from signals evoked in different gestational periods.

**Results:** The model revealed that the network’s intrinsic dynamics converge towards an endothelial preserve function. Additionally, using the DEGs of normal gestation as initial state has shown to preserve *eNOS* activity. Pathological perturbations, however, drove the system into a stable attractor characterized by the loss of eNOS activity. Crucially, two etiologically distinct pathways to the vascular dysfunction were identified: ***i***. an inflammatory NF-kB/TNF mediated which was triggered by either oxLDL or Inflammation inputs; and ***ii***.a “direct vascular-driven” pathway by oxidative stress. Despite their different routes, both pathways were characterized by inactive *AKT1*. Perturbation experiments confirmed that *AKT1* was the critical and main regulator of endothelial NOS function. While blocking upstream inflammation nodes did not prevent eNOS loss, the constitutive activation of AKT1 was consistent to maintain *eNOS* activity, superimposing any other pathological negative outcomes on vascular function.

**Conclusion:** This work raises AKT1 as a pivotal node to maintain endothelial NOS activity, even in the presence of oxidative and inflammation conditions. The model developed provides a mechanistic basis for the clinical heterogeneity of PE. At least two distinct molecular pathways were identified as “routes” that primed the network to PE’s attractors, i.e.: inactivation of eNOS. Nevertheless, AKT1 failure or inactive state was a *sine quo non* condition to drive PE. The findings imply that therapeutic approaches, including metformin, which aim to support or improve AKT1 and its downstream signaling, may be effective in promoting vascular health in pregnancies at high risk.

## 1. Introduction

Preeclampsia (PE) is a severe, pregnancy-specific hypertensive disorder that affects 2–8% of pregnancies worldwide and remains a leading cause of maternal and fetal morbidity and mortality (Ives et al. 2020). Clinically, it is characterized by new-onset hypertension after 20 weeks of gestation, frequently accompanied by systemic endothelial dysfunction impacting multiple organ systems (American College of Obstetricians and Gynecologists 2019). The pathogenesis of PE is understood to arise from placental dysfunction, driven by a complex interplay of inflammatory mediators, angiogenic imbalances, and metabolic changes (Phipps et al. 2019). Despite extensive research, the systems-level mechanisms that drive the transition from a healthy pregnancy to a stable pathological state are not fully elucidated.

One of the challenges in understanding PE is its dynamic nature. Normal placental development involves a precisely orchestrated temporal gene expression governed by several factors that implicate in biological processes of trophoblast differentiation, vascular remodeling, and metabolic adaptation (Turco et al. 2018; Vento-Tormo et al. 2018). Disruptions in these systems are hypothesized to be pivotal to PE pathogenesis (Staff et al. 2013; Leavey et al. 2016). However, much of the existing research relies on static, cross-sectional, studies that compare healthy and diseased tissues at a single time point (Dimitriadis et al. 2023). While these approaches have successfully identified many differentially expressed genes and proteins content (Mikheev et al. 2008; Sitras et al. 2009; Myers et al. 2013), they have the constraint of capturing the causal feedback loops and non-linear dynamics that drive disease progression (Gathiram and Moodley 2016). Therefore, these ‘snapshots’ capturing approaches can show which biomolecule is different, but not directly how the system behaves over time or how it responds to perturbations. To address this gap, systems biology can integrate different biological features into a time-aware model (Barabási, Gulbahce, and Loscalzo 2011). For this, Boolean network modeling provides a simple yet elegant scientific framework, allowing for the analysis of complex regulatory logic in systems where detailed kinetic parameters are unknown (Albert 2004; Wang, Saadatpour, and Albert 2012). By representing biomolecules as binary switches, these models can identify the stable attractor states of a network, which correspond to distinct cellular or physiological phenotypes (Fumia and Martins 2013). This method is particularly useful to investigate diseases like PE as states which arise from a module’s loss of regulation or between different modules and to identify the critical nodes and feedback loops that control the switch between health and disease.

This study employed an integrative computational strategy using database-derived PE static protein nodes which were mapped onto a time-point experimental transcriptomic from normal gestation. This intersected common genes, representing PE time-aware regulatory network, was used for diffusion analysis and Boolean dynamic modeling. Herein, the activation of *AKT1* by *INSR* or *EGFR* was shown to be fundamental to maintain endothelial synthesis of NO, even in the presence of a prior to gestational ongoing inflammation. Interestingly, *NF-kB* knockout was not able to impair the *eNOS* loss of function induced by inflammation. The diffusion analysis revealed that early pregnancy gene expression changes primarily diffused through inflammatory and stress response nodes such as *NFKB1* and *EPAS. in silico* perturbations, and heat diffusion propagation analysis provided that PECAM1 inactivation by oxLDL, and oxidative stress direct disruption of endothelial steady state response are independent perturbations which drive the system to PE phenotype. Nonetheless, independent of the perturbed biological signal previously mentioned, *AKT* has shown to be the major kinase that, when activated, is able to surpass any PE induce-signal and protect *eNOS* activity, hence endothelial integrity.

## 2. Methodology

### 2.1 Data Collection and Normal DEGs Identification

Time-series transcriptomic data from normal human placental tissues were retrieved from the Gene Expression Omnibus (GEO) under accession GSE9984 (Mikheev et al. 2008). The dataset comprised 12 samples representing three gestational periods: first trimester (45–59 days, n=4), second trimester (109–115 days, n=4), and term placentas (≥37 weeks, n=4). Differentially expressed genes (DEGs) were identified for each gestational transition (T1: Mid vs. Early; T2: Late vs. Mid; T3: Late vs. Early) using the GEO2R tool, which implements the limma package (Ritchie et al. 2015). To generate a broad candidate list for further network-level filtering, DEGs were defined using a threshold of | log_2_ fold change| > 0.58 and a nominal *P* < 0.05.

### 2.2 Proposed Workflow for Constructing the PE-Time-Aware Network

The work-frame used to develop an integrated temporal dynamic with static data of the pathology is represented in (Figure 1), and displayed as numbered step topics for clarity below.

**Figure 1.**
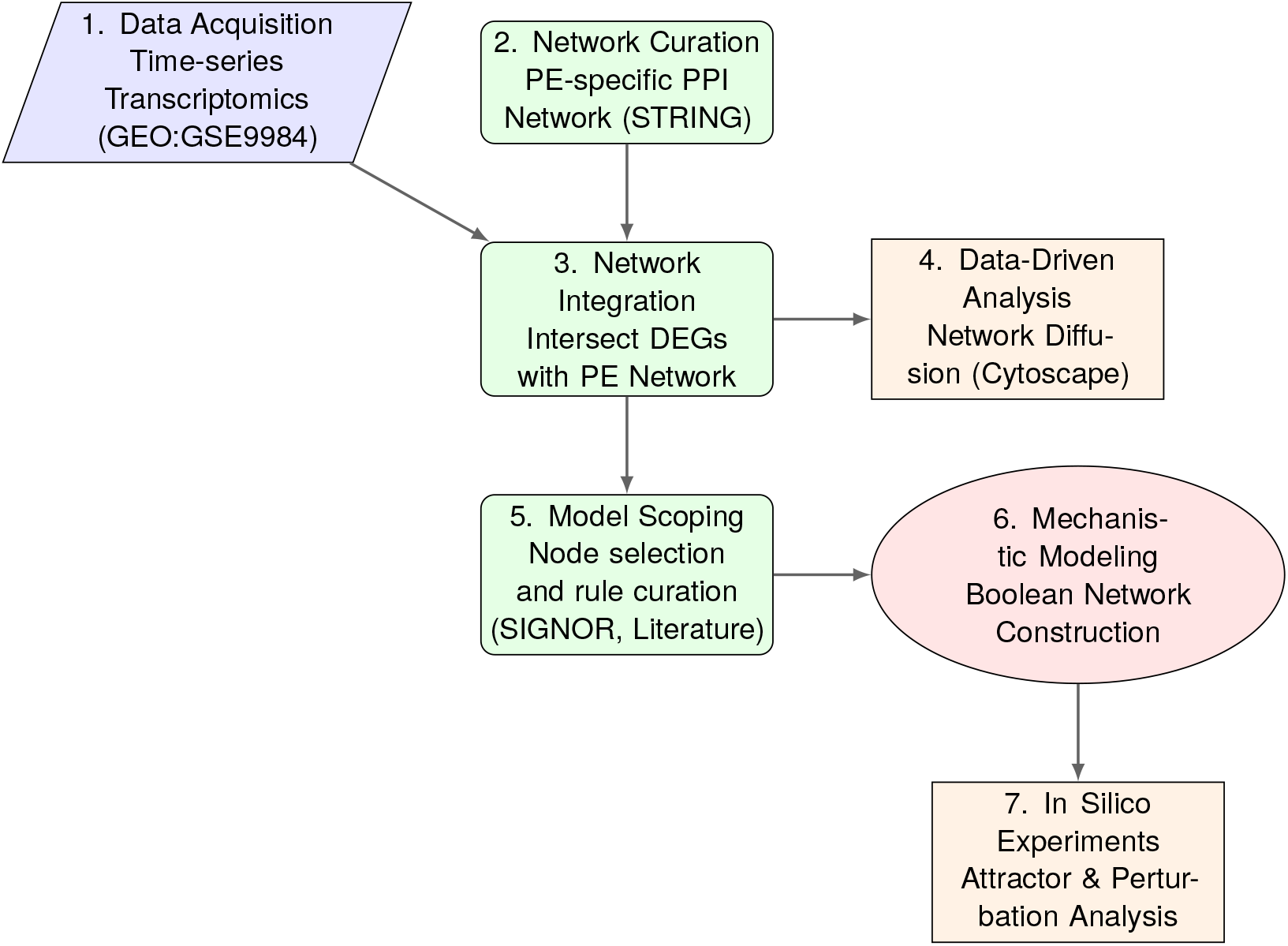
Integrative Systems Biology Framework for Modeling Preeclampsia Pathogenesis. The diagram illustrates the multi-step computational pipeline used in this study. The workflow begins with (1) acquiring time-series transcriptomic data and (2) curating a disease-specific protein network. These are (3) integrated to form a PE-Time-Aware network, which is subjected to two complementary analyses: (4) a data-driven network diffusion analysis to identify influential pathways, and a (5-6) mechanistic Boolean modeling approach to investigate causal logic. The Boolean model is then used for (7) in silico experiments, providing stable disease states, control points, and data-driven hypothesis generation for further experimental hypotheses on therapeutic targets.

**1. Preeclampsia Curated Network:** A high-confidence, PE-specific protein-protein interaction (PPI) network was constructed by querying the STRING database (v11.5) with the keyword “preeclampsia” (interaction score ≥ 0.9) (Szklarczyk et al. 2023). This curated network served as the disease-specific context.

**2. Identification of Temporally Regulated PE Genes:** The complete set of DEGs from all three gestational transitions was intersected with the curated PE network. This step identified a core set of genes that are both relevant to PE pathology and are dynamically regulated during normal pregnancy.

**3. Time-Specific Functional Enrichment:** The core set of genes from Step 2 was segregated into three lists based on the time point at which they were differentially expressed (T1, T2, or T3). Each of these three time-specific gene lists was then submitted to the Enrichr-KG platform for functional enrichment analysis (Kuleshov et al. 2016; Xie et al. 2021). This identified the key biological terms (e.g., pathways, diseases, transcription factors) associated with each phase of gestation.

**4. Iterative Network Expansion:** All enriched biological terms identified in Step 3 were compiled into a single, unified list. This list of terms was used to perform a second, iterative query on Enrichr-KG. This functional expansion step retrieved a comprehensive set of all genes and proteins strongly associated with the temporally dynamic processes relevant to PE. This final, expanded gene set constitutes the herein named “PE-Time-Aware Network”.

### 2.3 Data-Driven Analysis via Network Diffusion

To identify pathways that most influenced the network, heat diffusion analysis was performed on the PE-Time-Aware Network using the Diffusion algorithm in Cytoscape (Shannon et al. 2003; Otasek et al. 2019) using the default parameters. The initial DEGs from each transition (T1, T2, and T3) were used as distinct heat sources in three separate simulations to model the propagation of temporal signals through the functionally expanded network.

### 2.4 Construction of the Mechanistic Boolean Model

To investigate the causal logic of the system, a focused, mechanistic Boolean model was derived from the larger integrated network. A core set of 30 nodes was selected, representing key biological entities and signals across inflammatory, metabolic, endothelial, and hypoxia-response pathways. The directed regulatory relationships between these nodes were curated primarily from the SIGNOR 3.0 signaling network resource (Lo Surdo et al. 2023), with additional manual curation from literature to ensure biological relevance to PE.

Six external input nodes were defined to simulate exogenous conditions: ‘Inflammatory signal’, ‘Hypoxia signal’, ‘Oxidative stress’, and ‘oxLDL’ as pathological stimuli, and ‘Shear stress’ and ‘Insulin signal’ as homeostatic signals. The final model consists of these six inputs and 24 internal nodes.

### 2.5 Boolean Model and Simulation Analysis

The network was formalized as a Boolean system *B* = (*V, F*), where each node *v*_*i*_ ∈*V* can exist in a state of 1 (active/ON) or 0 (inactive/OFF). The logical update functions in *F* were formulated based on activating and inhibitory relationships from the curated network. Where multiple regulators converged on a single node without specified combinatorial logic, a majority-rule was used: a node activates if the sum of its activators exceeds the sum of its inhibitors. For interactions with known synergistic or conditional requirements, explicit AND/OR logic gates were implemented. The complete set of Boolean rules is provided in Supplementary Materials.

The attractors were identified by simulating state trajectories from 10,000 random initial conditions using a synchronous update scheme. Attractor basin sizes were estimated as the fraction of trajectories converging to each stable state. To identify critical control points, systematic *in silico* perturbations were performed by fixing individual nodes to a constant OFF (0) or ON (1) initial state and evaluating the shift in attractor basins. The significance of these shifts was assessed using Fisher’s exact test.

### 2.6 Software and Code Availability

Network construction and visualization were performed in Cytoscape (v3.9.1) (Shannon et al. 2003). The Boolean model was constructed and simulated using The Cell Collective platform (https://cellcollective.org) (Helikar et al. 2012), with further analysis performed using the PyBoolNet Python library (Klarner, Streck, and Siebert 2016). The model generated will be public available as soon as policies from ‘The Cell Collective’ are met.

## 3. Results

### 3.1 Summary of Temporal Placental Gene Expression from Uncomplicated Gestation

To establish a baseline of normal placental development, a time-series transcriptomic dataset (GSE9984) was analyzed. Pairwise comparisons between the first, second, and term trimesters identified 724 unique differentially expressed genes (DEGs), with the number increasing markedly across gestation (29 DEGs in T1, 139 in T2, and 656 in T3), indicating a major transcriptomic reorganization in the final trimester. Briefly, Table 1 raises conceptually extracted findings from normal gestational DEGs.

**Table 1.**
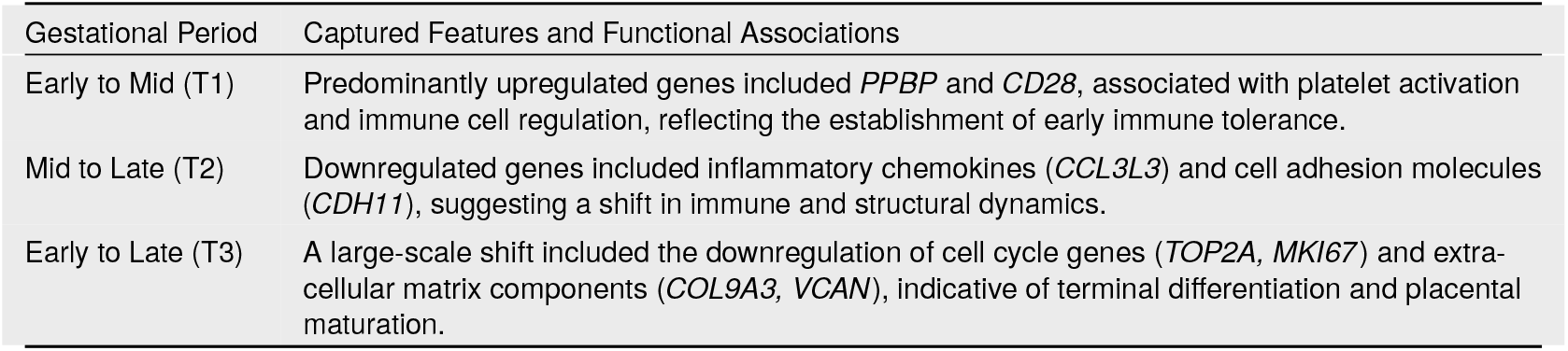
Simplified summary of differentially expressed genes across normal gestational periods.

Enrichment analysis of these DEGs revealed an overrepresentation of pathways critical to placental function, immune-metabolic modulation (*CD36, CD28, ACACA, ABCG1*), cell signaling (*BMP5, ACVR2B*). Notably, distinct inflammatory signatures were observed at each stage, with *TNF* and *NF-kB* signaling prominent in early gestation, transitioning to IL-6/JAK/STAT signaling mid-gestation, and finally to complement and coagulation cascades at term.

### 3.2 Implementation of an Integrated, Multi-layered and ‘Time-Aware’ Preeclampsia Network

To investigate potential nodes that were involved in PE onset and progression across gestation, a high-confidence, static PE network of 459 proteins was first extracted from the STRING database (Supplementary Figure S1). This static disease network was then intersected with the 724 temporal DEGs from the normal placental time-point dataset, as previously identified. The common time-dependent nodes were enriched using a comprehensive multiomics approach. Terms and biological features retrieved in each time-point can be seen in Table 2.

**Table 2.**
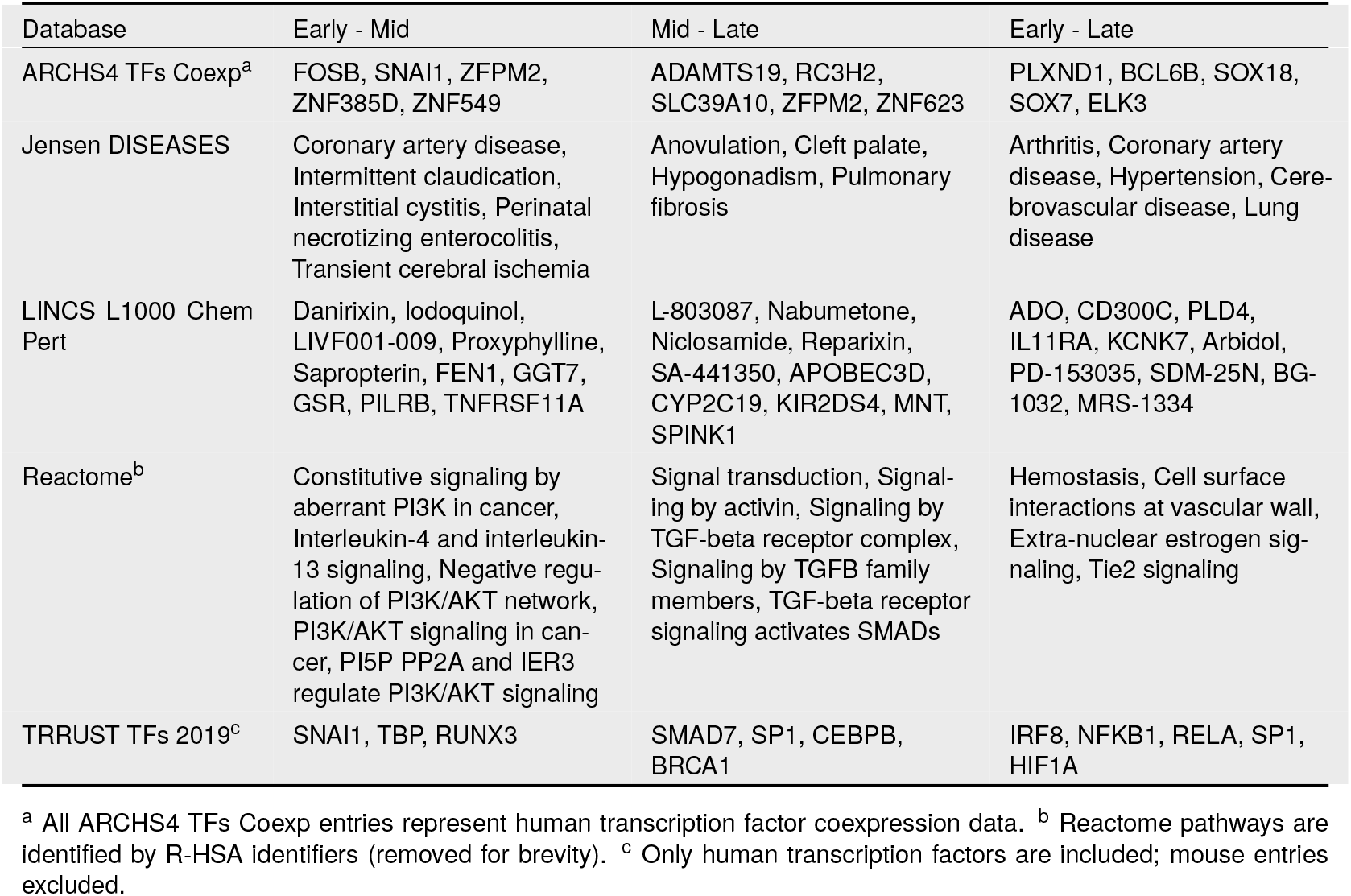
Comparison of enriched biological terms across different databases by time points from the intersected nodes, in which PE retrieved protein-protein interactions were mapped onto DEGs from experimental dataset of time dependent normal gestation development.

A second enrichment was performed, according to the pipeline in here proposed, where all genes retrieved from the first step were used to build a final “PE-Time-Aware Network” that is represented in Figure 2. This integrated network was the foundation for both data-driven and mechanistic modeling, containing the PE-relevant nodes by which expression could be dynamically regulated in the disease onset and progression.

**Figure 2.**
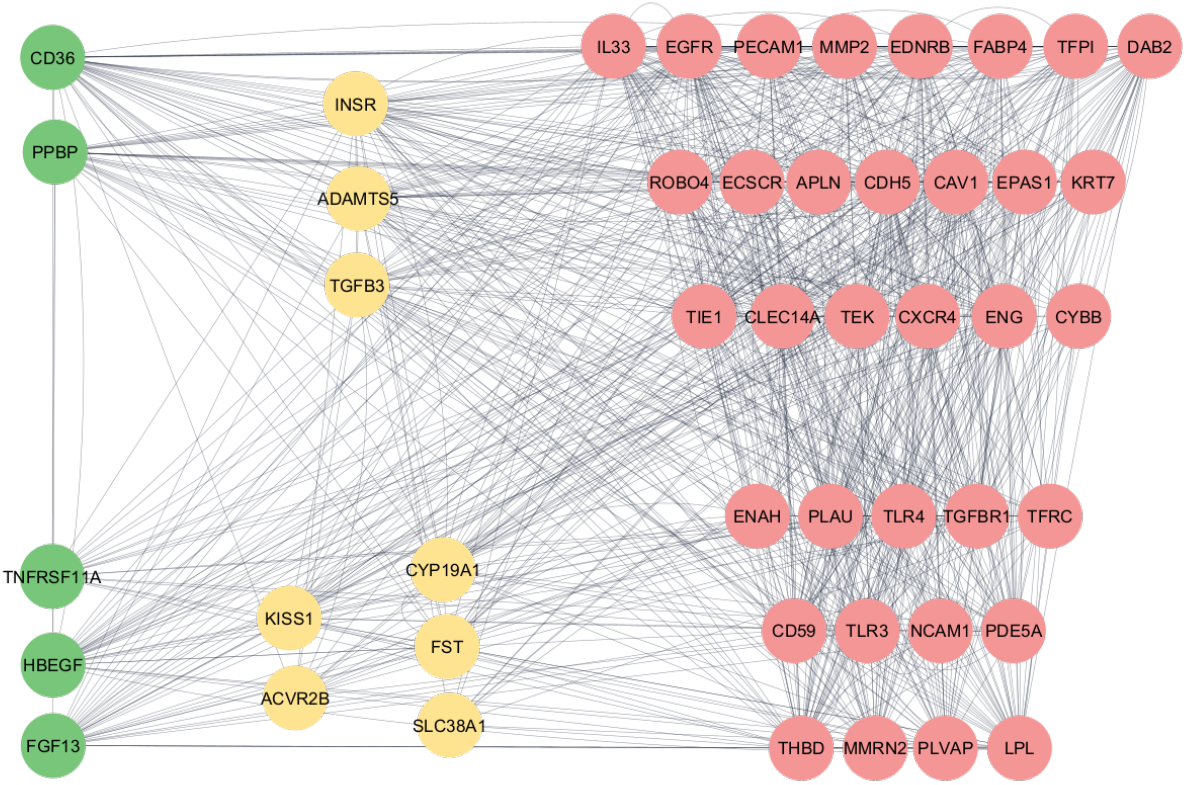
The Integrated PE-Time-Aware Network. The network visualizes functional associations among genes linked to both preeclampsia and differential expression during normal gestation. Nodes are colored by the gestational stage of their peak expression: T1 (early, green), T2 (mid, yellow), and T3 (term, red). Edges represent known or predicted functional relationships from the GeneMANIA database. The dense interconnectivity shows that genes from different temporal stages form a functionally integrated system. This network development followed the framework described in the Methods sections.

This second round of enrichment analysis provided highly connected hubs associated with specific pathological phenotypes, such as Atherosclerosis, Myocardial Infarction, and Coronary Artery Disease. *CCL2, MYDGF, JCAD*, and *ENPEP* were representative signature nodes from the different sets of genes related to each of those pathologies. The Supplementary Table S2 presents the full data of the hubs and the genes involved in each of the identified pathological phenotypes.

### 3.3 Analysis of Temporal Cascade of PE Time-Aware by Diffusion Algorithm

Network propagation analysis was performed on the PE-Time-Aware Network to identify which nodes were most proximal to the temporally expressed seed genes at each time point. Notably, independently of seed genes representing the temporal propagation, Insulin Receptors downstream nodes were identified as significantly influenced targets. In Early to Mid-Gestation transition (Figure 3a), the inflammatory and metabolic seed genes (e.g., *CD36, PPBP*) propagated through nodes related to cellular stress response and growth factor signaling. A striking influence on *EGFR*-associated genes was seen. Additionally, the top-ranked genes included the autophagy regulator *TFEB* and the receptor tyrosine kinase *ERBB4* (Supplementary Table S3). In the Mid to Late-Gestation (Figure 3b), genes involved in regulation of placental development (*KISS1, GATA2*), that propagated their signal to members of the TGF-beta family, such as *GDF11*. Less pronounced inflammatory nodes could be noticed at this stage, being metabolic and cellular oxygen handling genes among the most diffused ones. Finally, in the Early to Term comparison (Supplementary Figure 3c), the wide spectrum of endothelial and vascular seed genes (e.g., *KLF2, ANGPT1*) propagated their signal through kinase *SRC* and the metabolic/inflammatory regulators, such as *PPARG, FABP4, CCXR4, AGTR1, IL33*. This may represent a potential dysfunctional axis involved in the endothelial damage generated by a systemic unresolved inflammation.

**Figure 3.**
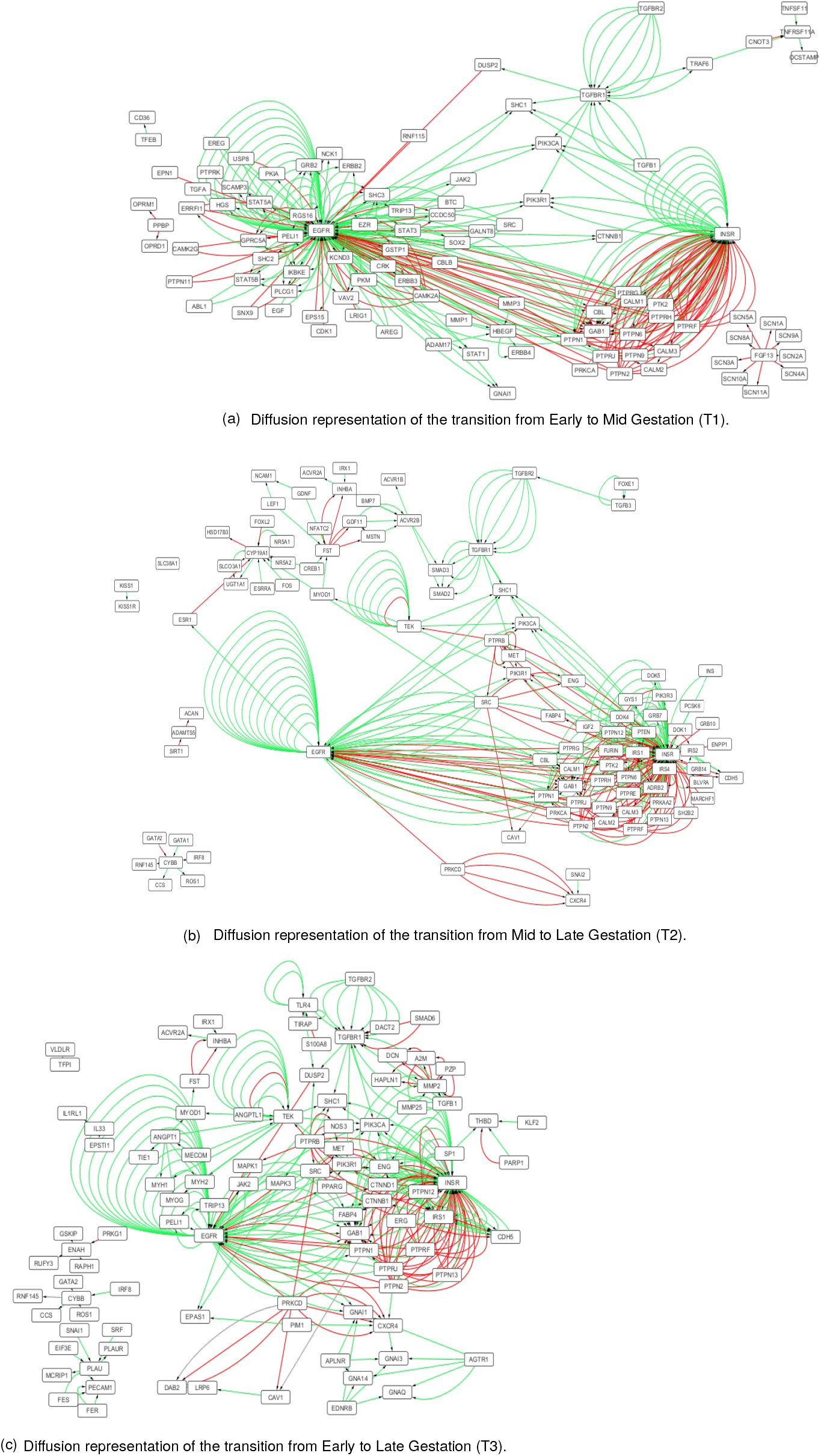
Network Heat Propagation/Diffusion obtained by using the time-specific deferentially expressed genes from mid vs. early, late vs. mid, and late vs. early comparisons as seeds to three independent diffusion simulations. This approach highlights potential genes and biological processes that are most influenced during disease progression

### 3.4 Boolean Model Network Exhibited Intrinsic Health Endothelial Attractor

The obtained Dynamic PE-Aware Model and its external signals are represented by Figure 4a. Analysis of the Boolean model’s logic revealed a naturally occurring behavior characterized by a “healthy” attractor which preserves *eNOS* activity, hence the endothelial function. In the absence of all external signals and all nodes in set to inactive initial condition, a dynamic oscillating angiogenic module among *sFLT, CDH5, HIF* and *VEGFA* marked the “healthy” normal attractor (Figure 4b). This physiological loop may indicate a dynamic fine-tuning regulation of vascular integrity as a response to small local oxygen fluctuations. Additionally, when the positive “homeostatic” stimuli Shear Stress and Insulin were applied on this network, the *eNOS* preserved function was also seen with the same cycling module previously described (Figures 4c) which independently confirmed the robustness of model deployment.

**Figure 4.**
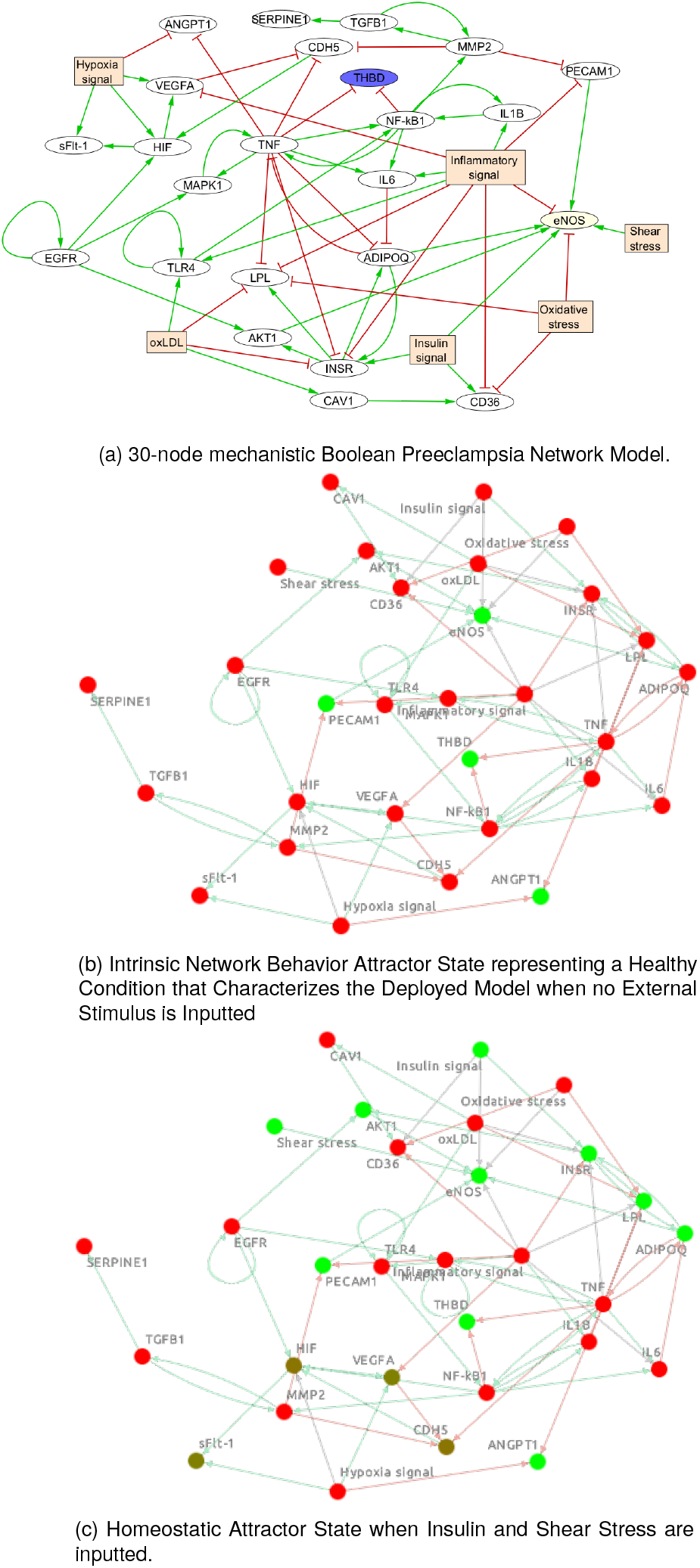
Model Representation and The “Homeostatic” Behavior Stable State of the Preeclampsia Time-Aware Network. (a) The illustration depicts the Boolean Dynamic Model representing the PE-Time-Aware. Directed Edge relationships between nodes are shown either by arrow (→) in the color green, which represents the rule “activates”, or the arrow red, meaning an inactivation property the source to the target. (b) Network Intrinsic Behavior Attractor State without external perturbations or inputs. (c) Homeostatic Attractor where no Pathological Stimuli are present, but Insulin and Shear Stress are inputted as positive metabolic and vascular external stimuli. In both (b) and (c) conditions a fine-tuned loop among *CDH5, sFLT, VEGFA, and HIF* cycling attractor (nodes highlighted darker in (c) only). All nodes were set to be inactive as the initial state.

The model predicted that different pathological stimuli can lead to endothelial dysfunction either by direct inhibition of *eNOS* when the Oxidative Stress was inputted (Figure 5a) or by *PECAM-1* inactivation caused by oxLDL external perturbation (Figure 5b). Therefore, Figure 5 shows that Oxidative Stress input promoted a direct *eNOS* inactivation, which was reversible upon removal of stimulus, despite maintaining *PECAM1* activity. On the contrary, oxLDL inactivated *eNOS* irreversibly, and this effect was attributed to uttermost *PECAM1* inactivation. Notably, in both conditions the Insulin and Shear Stress were absent in the simulations.

**Figure 5.**
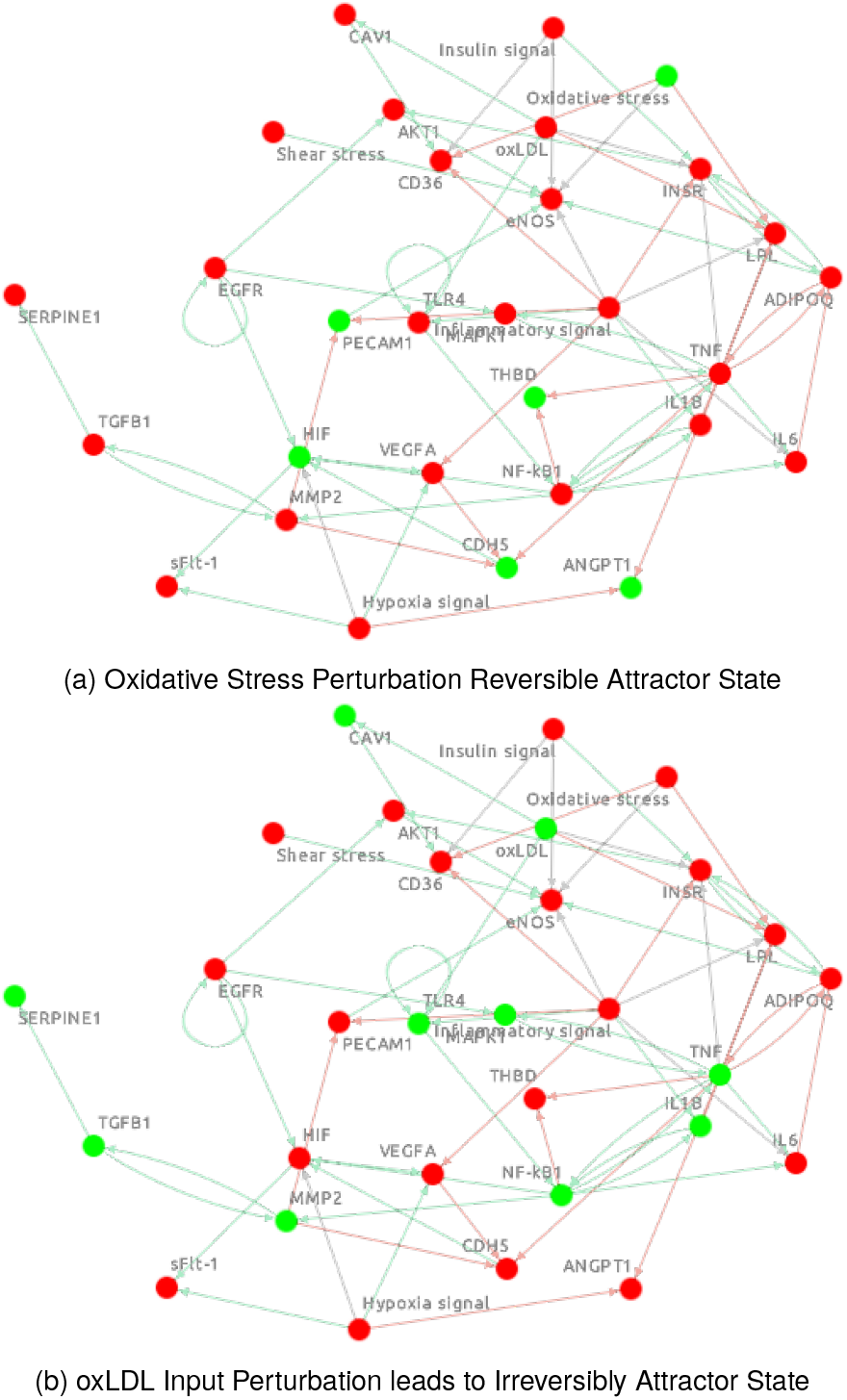
Boolean Network Behavior Dynamics During Different Stimuli Perturbations. The figure shows the state’s attractors that the Boolean network converges upon (a) Oxidative Stress and (b) oxLDL independently solely inputted. Node colors indicate the state: green (ON/active) and red (OFF/inactive).

### 3.5 Endothelial Dysfunction in Preeclampsia Is Primarily Driven by Metabolic Disruption of INSR/AKT Signaling Rather than Direct Inflammatory Activation Only

Next, the model was used to test the causal drivers of endothelial dysfunction, a clinical hallmark of PE which the outcome inhere was represented by the activity of *eNOS. In silico* perturbation experiments revealed that the integrity of the metabolic signaling axis, not the absence of inflammation, was the critical determinant of endothelial health (Figure 6). Knockdown of the main inflammatory ‘bottleneck’ signaling node *NFKB1* was not enough to rescue *eNOS* activity either with chronic (the model begins the simulation with inflammatory stimuli activated, Figure 6c) or acute inflammation from the PEA state’s fate. The constitutive activation of *INSR* or *AKT1* fully restored eNOS function, even under maximal inflammatory stimuli input (Figure 6d and, respectively). These results provide a hypothesis that the failure of the insulin/AKT1 pathway is the main mechanism that drives the loss of endothelial homeostasis in this model.

**Figure 6.**
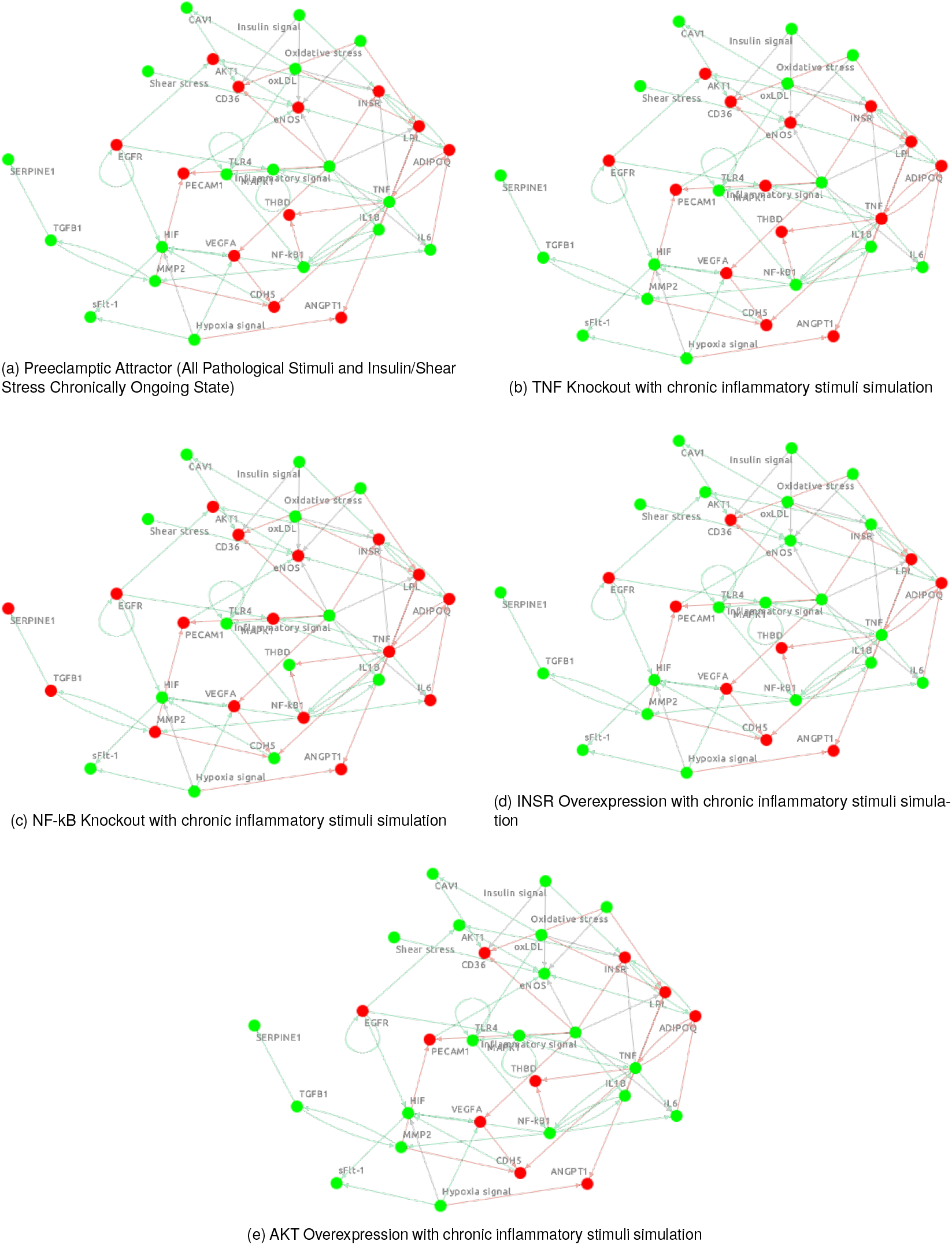
Metabolic Signaling Activation is the Major Determinant to Endothelial Function, Even in the Presence of Inflammatory Signals. The figure displays the stable states (attractors) of the Boolean network under different perturbation scenarios. (a) The Preeclamptic Attractor State (PEA) shows a fully active inflammatory module and complete suppression of endothelial function (*eNOS* is OFF, in red). Knockout of (b) *TNF* or (c) *NF-kB* does not restore endothelial function, as seen by *eNOS* maintaining OFF state in both independent simulations. Constitutive overexpression of (d) *INSR* or (e) *AKT1* was able to rescue endothelial function (*eNOS* active in green) despite of all persistent pathological stimuli activated.

### 3.6 Uncomplicated Gestational DEGs Show Preserved eNOS activity and AKT or EGFR inactivation, but not INSR, are the Major Drivers Nodes of PE

Further *in silico* perturbations were conducted by setting the initial state of those upregulated genes collected from normal gestational to be activated. Hence, the initial states of *ANGPT1, CAV1, CDH5, EGRF, HIF, INSR, LPL, PECAM1, SERPINE1, TGFB1, THBD, TLR4, and TNF* was set to be active and the remaining inactive before running the model. Interestingly, the stimuli initial conditions, i.e.: either none or all active inputs, did not affect the sustained activation of *eNOS* (Figure 7a). To understand this network behavioral in this condition, one-by-one of the upregulated genes was set to inactive while others were held at the activated initial condition, independently. As seen in Figure 7b, EGRF was the only gene that when inactivated led to *eNOS* loss of function maintaining all-stressors fully activated. AKT Knockout (Figure 7d, but not INSR (Figure 7c), led to loss of *eNOS* function seen in upregulated genes initial setup with all deleterious stimuli on.

**Figure 7.**
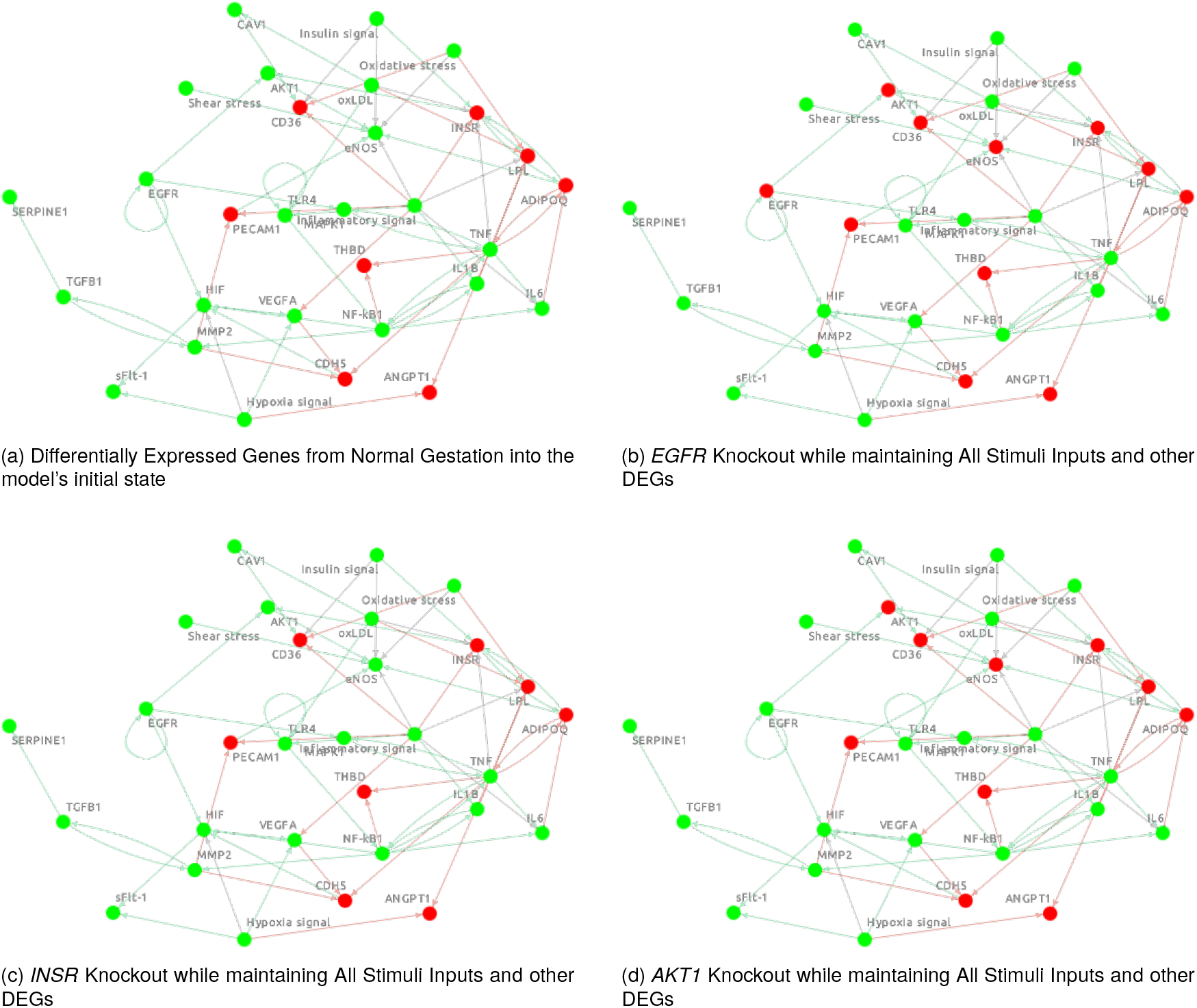
Differentially Expressed Genes Associated with Uncomplicated Gestational as a proof-of-concept initial signature to preserve *eNOS* and vascular function. *AKT1* and *EGFR* key nodes have shown to be Critical for Endothelial Health under Different Pathological Stimuli. The figure displays the state attractors based on normal gestational gene expression. Node colors indicate the state: green (ON/active), red (OFF/inactive). (a) The model was run with the initial state set to key normal gestational genes and all six external stimuli active. Notably, endothelial function is maintained (i.e.: active *eNOS*) after reaching the attractor state, demonstrating the system’s robustness.(b) Among Knockout of *EGFR* is unique to cause *eNOS* OFF. (c) In contrast, the knockout of *INSR* has no additional effect on the already stressed system, as its pro-survival signaling pathway is suppressed by the inflammatory environment. (d) The direct knockout of *AKT1* invariably leads to the loss of *eNOS* function, confirming its role as the “switcher” for endothelial health.

## 4. Discussion

This study used an integrated systems biology approach, combining multi-level biological data retrieved from time-point transcriptomic analysis to understand the dynamic molecular mechanisms associated with preeclampsia onset and progression (PE). A robust framework was developed and a data-driven hypothesis that PE uttermost could be a metabolic failure where molecular events would converge to an irreversible inactivation of the AKT1. This latter process would void the pass-through signal to *eNOS* and, consequently, lead to the loss of endothelial function. The Boolean model developed here provides a mechanistic explanation for the complex clinical features of PE, which has traditionally been viewed as a single pathway failure. Instead, the present work suggests that PE is characterized by at least three distinct pathways. A direct, inflammatory and a central metabolic pathway.

This “metabolic failure” hypothesis provides a mechanistic framework that elegantly synthesizes numerous clinical observations. It is well-established that metabolic syndrome, obesity, and pre-existing insulin resistance are major risk factors for PE (Karumanchi et al. 2005; Weissgerber and Mudd 2015). A causal explanation for this correlation can be drawn from the fact that pregnancies in metabolically compromised conditions may reduce allostatic load capacity to handle drastic stress during pregnancy (McEwen 2007; Hruby and Hu 2016). Especially, oxygen pressures fluctuations, immune-inflammatory high-and-lows put extra ‘pressure’ on the high all-time long demanded insulin downstream responses.

The results found also shed light on the extent severity seen in PE cases. That is in agreement to the clinical endpoint of endothelial dysfunction (Powe, Levine, and Karumanchi 2011; Mol et al. 2016). A direct vascular subtype may be driven by oxidative stress, a well established eNOS uncoupling (Santillo et al. 2015), while an inflammatory” subtype may be driven by insults like ‘oxLDL’ that dismantle the adaptive AKT1 pathway via inflammatory suppression of insulin signaling (Zhu et al. 2016; Bilodeau 2018). This explains how different patients with different underlying vulnerabilities can arrive at a similar clinical presentation (Leavey et al. 2016).

Strategies focused solely on broad anti-inflammatory action may be less effective than those aimed at preserving metabolic resilience (Palei et al. 2013; Cornelius, Amaral, and Wallace 2018). The model predicts that therapies capable of preserving the downstream AKT effective effectors could shield the endothelium from damage, representing a more targeted and effective approach (Zeng et al. 2017). This aligns with emerging evidence for the potential use of metformin, an insulin-sensitizing drug, in reducing the risk of preeclampsia (Romero et al. 2017; Brownfoot et al. 2021). By identifying a mechanistic link between metabolic resilience and vascular health, this work provides a strong rationale for exploring such metabolically targeted interventions and drug-repurposing strategies.

Systems biology and translational medicine require increasing integration of research. A crucial, practical question bridging bench and bedside research is whether preeclampsia is an acute hypertensive disorder or a latent, asymptomatic condition progressing naturally to hypertension, speeded up by gestational physiological stress exceeding the biological system’s stability threshold and triggering pathology. This study suggests a more significant role for gestational or pre-existing diabetes in preeclampsia pathogenesis than initially assumed, prompting metformin repositioning as a potential preeclampsia treatment. The clinical presentation’s broad spectrum of symptoms and outcomes requires consideration of all factors. This research emphasizes the promising potential of drug re-purposing, a faster and more efficient alternative to new drug development, especially for many at-risk conditions.

### 4.1 Limitations

The limitations of the study involve the following aspects: First, the Boolean abstraction necessarily simplifies continuous biological processes; while this allows for the analysis of large network logic, it may miss more subtle, dose-dependent effects. Second, the model currently lacks explicit representation of the maternal-fetal interface or systemic maternal cardiovascular factors, focusing instead on the core placental cellular network. Third, the network topology, while curated from high-confidence sources, may not fully capture all pregnancy-specific interactions. Finally, all findings are computational and require experimental verification.

### 4.2 Future Directions

Future work should prioritize the validation of this model’s predictions in larger, prospective longitudinal cohorts. Concurrently, experimental validation is essential, using in vitro models such as placental explants to functionally validate the predicted roles of key hub genes like *AKT1* and confirm the dynamics of the “metabolic-inflammatory switcher”. The computational framework itself can be enhanced by advancing to more sophisticated formalisms, such as probabilistic or hybrid models, to increase predictive accuracy. Ultimately, this refined model can be leveraged for in silico therapeutic screening to identify and prioritize interventions that either “teach” the body to do its job very well or create a system-level metabolic shield during the critical window of early gestation.

## 5. Conclusion

By integrating time-series transcriptomics with both mechanistic and data-driven network modeling, this study provides a dynamic, systems-level framework for understanding preeclampsia pathogenesis. The work has provided a time view multiomics work-frame that comes up with a Boolean Model that reveals a switch governed by the antagonism between metabolic health and inflammatory stress, and raises forward whether the novel hypothesis that the failure of the AKT1 “Switcher” signaling axis is the primary driver of endothelial dysfunction. The identification of a critical transition window and the protective role of metabolic resilience offer a new foundation for revisiting old known strategies, especially considering a deeper knowledge in similar conditions that does not involve pregnancy.

## Data Availability

The model generated will be public available as soon as policies from The Cell Collective are met.

## Acknowledgment

The author gratefully acknowledges the Federal University of Fronteira Sul for the support and the opportunity to dedicate full-time efforts toward the development and completion of the present study.

## Data Availability

The model and the complete set of Boolean rules are available in the Supplementary Material. The source model used in this study will be publicly available on The Cell Collective platform following peer review and publication of this article.

## Generative AI and Foundation Models Disclaimer

The author declares the use of the state-of-art large language model from Google AI Studio *(gemini-2*.*5-pro-preview-06-05 with default sampling settings*) for assistance in Python coding/linting, English language readability, and LaTeX formatting to improve the professional styles’ quality of the manuscript. Prompting engineering by branching techniques (long context awareness environment), zero shot prompts and *persona* specification to model were used. MCP was used to give the model access to authors’ workspace environment to improve context, history logs and output performance of the model. The data output, chat history and prompts are entirely registered and stored in the author’s personal database and backup scheme. The conception, development and all scientific rationale are solely derived from author’s intensive study and work. To the author’s best knowledge, no Generative Model exists that is able to hypothesize and originally provide the comprehensive formed scientific narrative and *in silico* designs herein presented. The author takes full responsibility for all content, including all the original results, computed experiments, scientific interpretation, conclusions drawn, and final manuscript.

## Supplementary Material

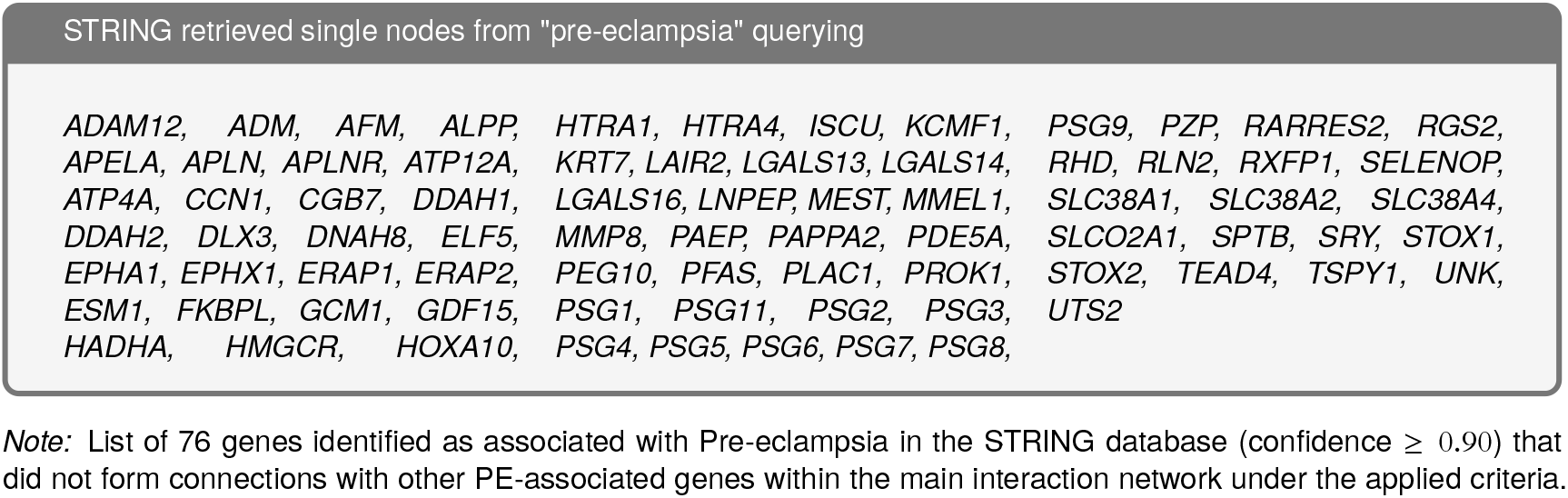

**Figure S1.**
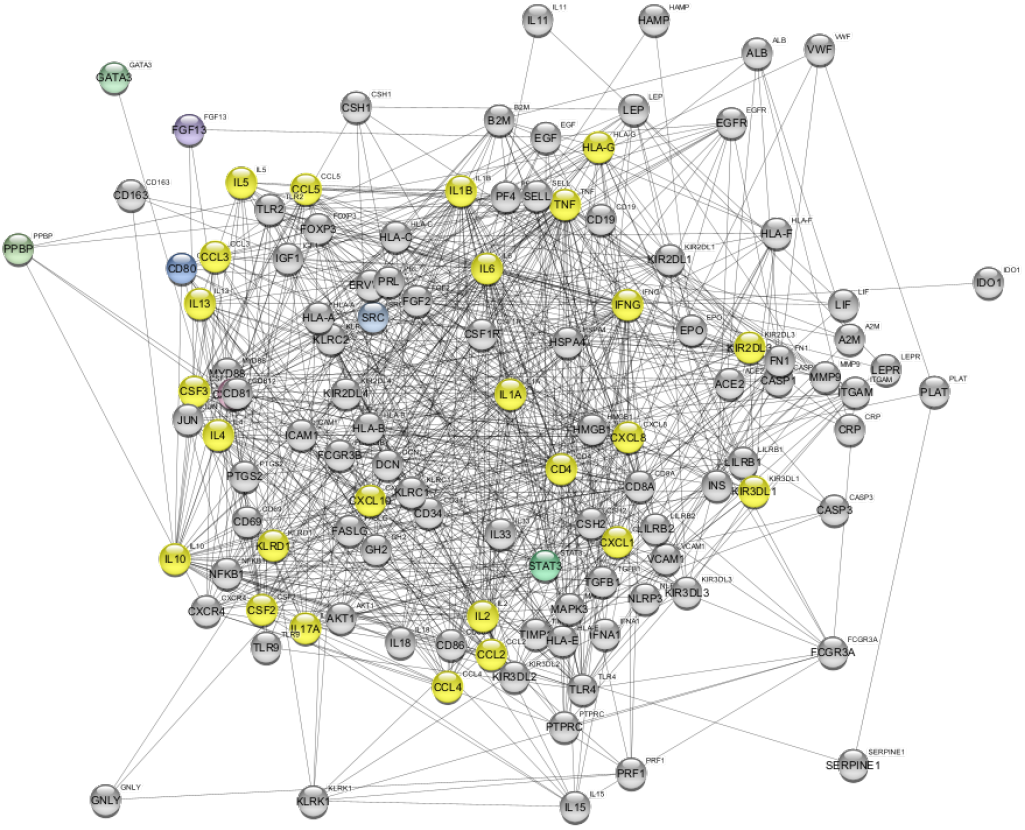
Preeclampsia-associated protein–protein interaction (PPI) network derived from STRING database analysis. The network displays high-confidence interactions among proteins curated for their relevance to preeclampsia (PE). Nodes highlighted in yellow represent the top 25 hub proteins identified by Maximal Clique Centrality (MCC) analysis, suggesting their central regulatory roles within the network architecture. Node color gradients denote functional grouping or biological annotation, while edges represent experimentally supported or strongly predicted interactions (confidence score ≥0.9). This hub-centric topology underscores key molecular regulators that may coordinate the complex pathophysiology of PE.

**Table S1.**
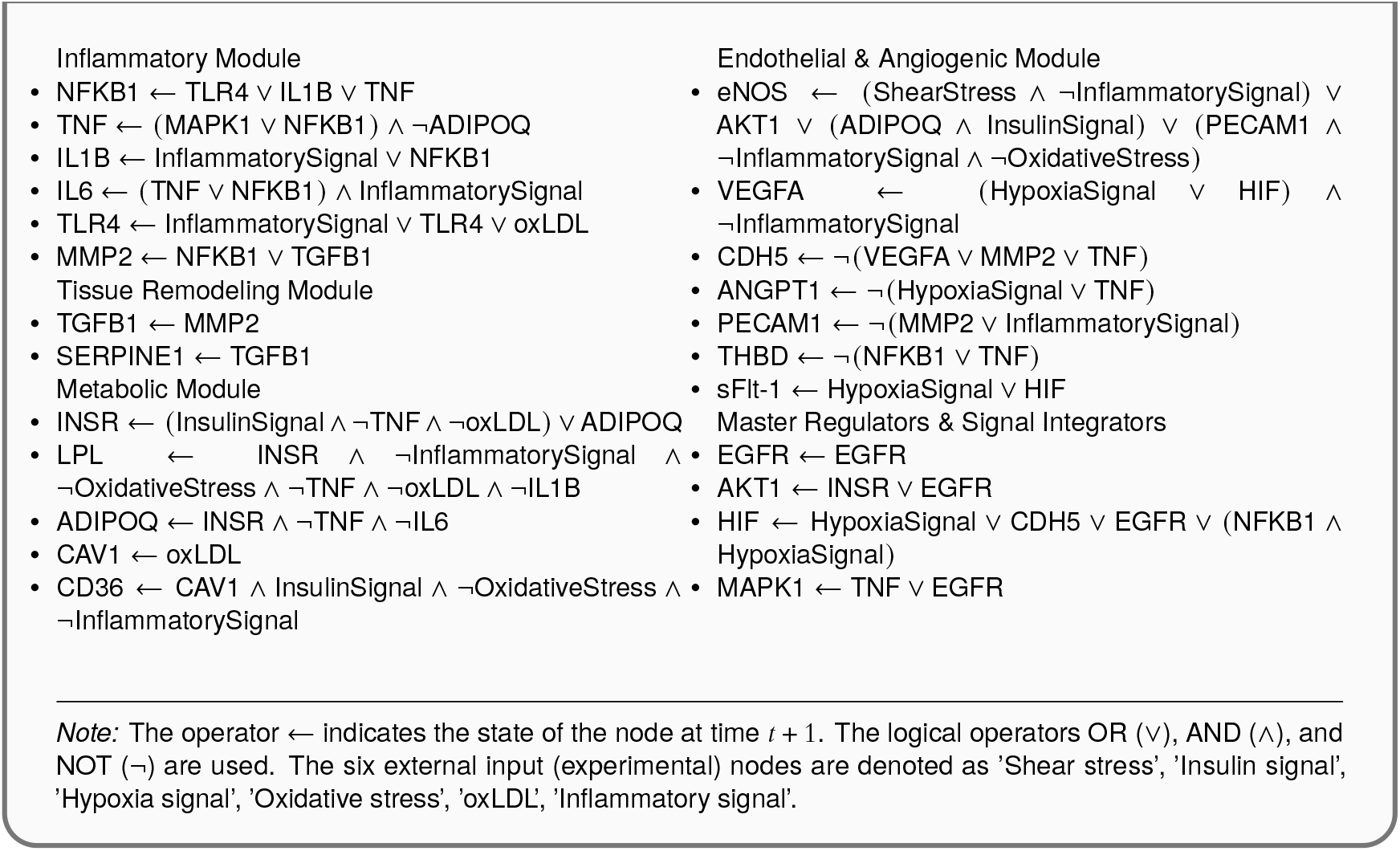
Boolean Network Logical Rules.

**Table S2.**
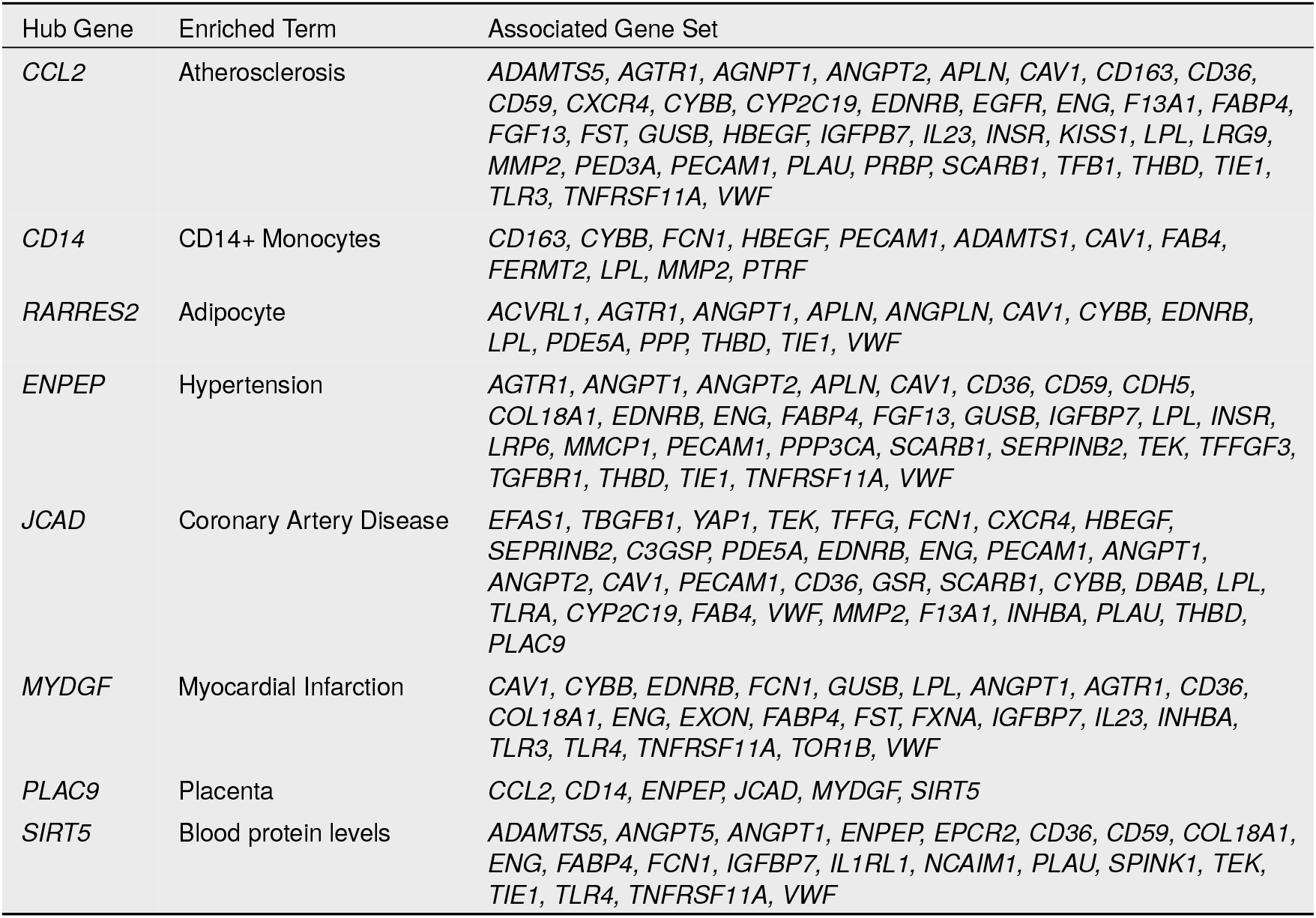
Hub genes identified from the intersected network, their associated enriched terms (phenotypes), and the corresponding gene sets.

**Table S3.**
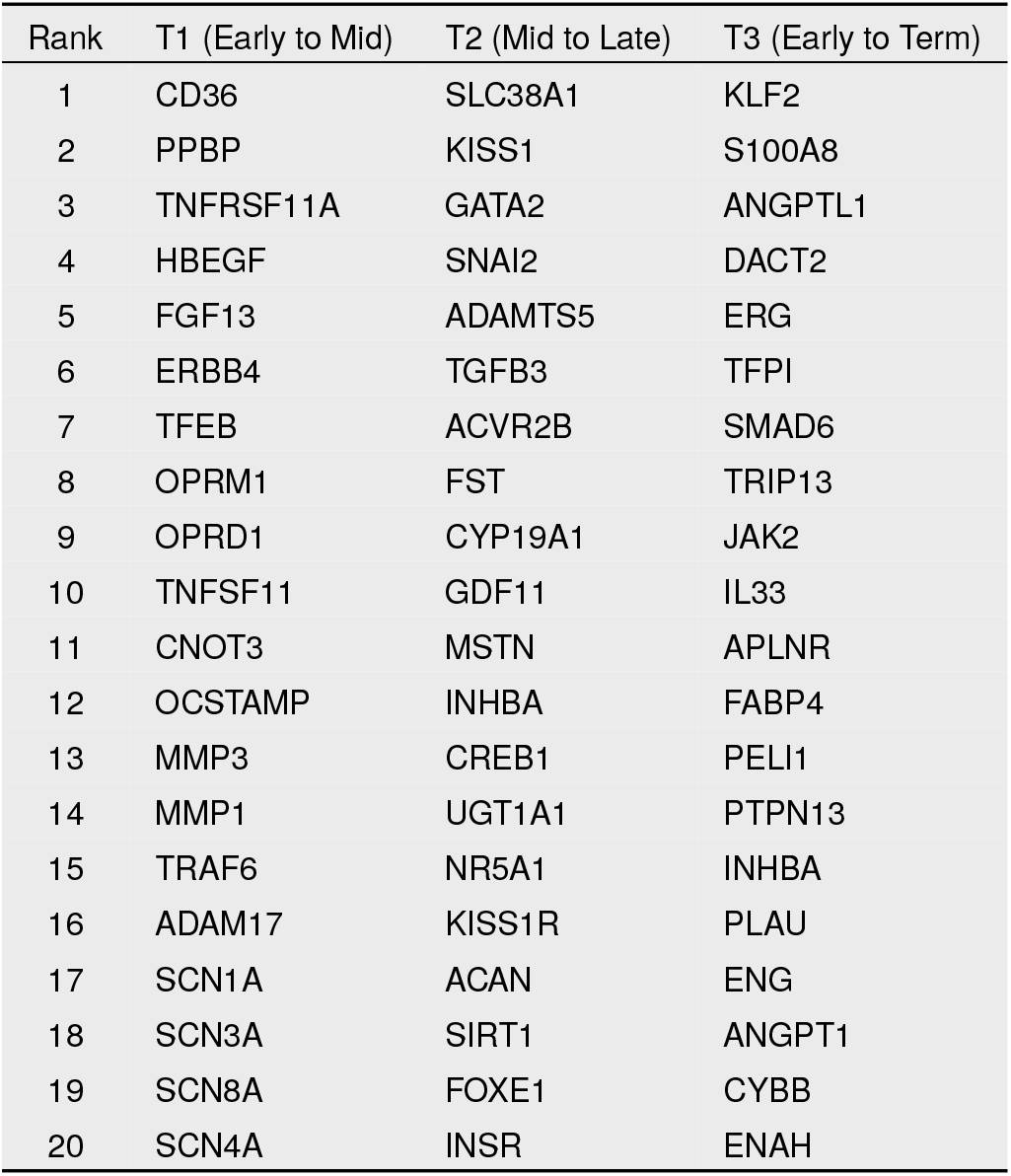
Top 20 Ranked Genes from Network Diffusion Analysis at Each Gestational Transition. The table lists the most influential genes based on their final heat score after simulations were seeded with temporally expressed DEGs. Genes that were part of the initial seed set for each run are shown in bold, allowing for the identification of the key non-seed nodes that are most functionally proximal to the temporal signals.

**Table S4.** In Silico Perturbations Reveal Metabolic Control of eNOS Activity. The table shows the final stable state of *eNOS* under different perturbation scenarios, all simulated in the presence of strong, persistent pathological stimuli (Inflammation, Hypoxia, oxLDL, and Oxidative Stress).

| Intervention Type | Perturbation | Final eNOS State |
| --- | --- | --- |
| Baseline (Full Pathological Stimuli) |  | 0% (OFF) |
| Anti-Inflammatory "Mimicking" Interventions |  |  |
|  | *NFKB1* Knockout (OFF) | 0% (OFF) |
|  | *TNF* Knockout (OFF) | 0% (OFF) |
| Metabolic/Homeostatic "Mimicking" Interventions |  |  |
|  | *INSR* Overexpression (ON) | 100% (ON) |
|  | *AKT1* Overexpression (ON) | 100% (ON) |

## References

Albert, Réka. 2004. Boolean modeling of genetic regulatory networks. Chaos: An Interdisciplinary Journal of Nonlinear Science 14 (4): 1038–1049. 10.1063/1.1414882.

American College of Obstetricians and Gynecologists. 2019. Gestational hypertension and preeclampsia: acog practice bulletin, number 222. Obstetrics and Gynecology 133 (6): e1–e25. 10.1097/AOG.0000000000003281.

Barabási, Albert-László, Natali Gulbahce, and Joseph Loscalzo. 2011. Network medicine: a network-based approach to human disease. Nature Reviews Genetics 12 (1): 56–68. 10.1038/nrg2918.

Bilodeau, Josee-Francois. 2018. Oxidized ldl and preeclampsia: linking placental ischemia to maternal vascular dysfunction. Hypertension in Pregnancy 37 (2): 93–103. 10.1080/10641955.2018.1441793.

Brownfoot, Fiona C, Roxanne Hastie, Natalie J Hannan, Ping Cannon, Laura Tuohey, Laura J Parry, Sevvandi Senadheera, Sebastian E Illanes, Tu’uhevaha J Kaitu’u-Lino, and Stephen Tong. 2021. Metformin as a prevention and treatment for preeclampsia: effects on soluble fms-like tyrosine kinase 1 and soluble endoglin secretion and endothelial dysfunction. American Journal of Obstetrics and Gynecology 214 (3): 356.e1–356.e15. 10.1016/j.ajog.2015.12.019.

Cornelius, Denise C, Lorena M Amaral, and Kedra Wallace. 2018. Novel therapeutic targets for preeclampsia. International Journal of Molecular Sciences 19 (3): 863. 10.3390/ijms19030863.

Dimitriadis, Evdokia, Daniel L Rolnik, Wenda Zhou, Guadalupe Estrada-Gutierrez, Keiichi Koga, Ryan Paul Vieira Francisco, Clare Whitehead, Jon Hyett, Fabricio da Silva Costa, Kypros Nicolaides, et al. 2023. Pre-eclampsia. Nature Reviews Disease Primers 9 (1): 8. 10.1038/s41572-023-00417-6.

Fumia, Herman F, and Marcelo L Martins. 2013. Boolean network model for cancer pathways: predicting carcinogenesis and targeted therapy outcomes. PLoS One 8 (7): e69008. 10.1371/journal.pone.0069008.

Gathiram, P, and J Moodley. 2016. Preeclampsia: a review of the pathophysiology and the role of the renin-angiotensin-aldosterone system. International Journal of Gynaecology and Obstetrics 134 (1): 1–7.

Helikar, Tomáš, Bryan Kowal, Sean McClenathan, Mitchell Bruckner, Thaine Rowley, Alex Madrahimov, Ben Wicks, Manish Shrestha, Kahani Limbu, and Jim A Rogers. 2012. The cell collective: toward an open and collaborative approach to systems biology. BMC Systems Biology 6 (1): 1–14. 10.1186/1752-0509-6-96.

Hruby, Adela, and Frank B Hu. 2016. The metabolic syndrome: a global public health problem and a new definition. Journal of Allergy and Clinical Immunology 137 (6): 1710–1712. 10.1016/j.jaci.2016.01.035.

Ives, C William, Rachel Sinkey, Indranee Rajapreyar, Alan TN Tita, and Suzanne Oparil. 2020. Preeclampsia—pathophysiology and clinical presentations: jacc state-of-the-art review. Journal of the American College of Cardiology 76 (14): 1690–1702. 10.1016/j.jacc.2020.08.014.

Karumanchi, S Ananth, Sharon E Maynard, Isaac E Stillman, Franklin H Epstein, and Vikas P Sukhatme. 2005. Preeclampsia: a renal perspective. Kidney International 67 (6): 2101–2113. 10.1111/j.1523-1755.2005.00316.x.

Klarner, Hannes, Adam Streck, and Heike Siebert. 2016. Pyboolnet: a python package for the generation, analysis and visualization of boolean networks. Bioinformatics 33 (5): 770–772. 10.1093/bioinformatics/btw682.

Kuleshov, Maxim V, Matthew R Jones, Andrew D Rouillard, Nicolas F Fernandez, Qiaonan Duan, Zichen Wang, Simon Koplev, et al. 2016. Enrichr: a comprehensive gene set enrichment analysis web server 2016 update. Nucleic Acids Research 44 (W1): W90–W97. 10.1093/nar/gkw377.

Leavey, Katherine, Stephen J Benton, David Grynspan, John CP Kingdom, Shannon A Bainbridge, and Brian J Cox. 2016. Unsupervised placental gene expression profiling identifies clinically relevant subclasses of human preeclampsia. Hypertension 68 (1): 137–147. 10.1161/HYPERTENSIONAHA.116.07293.

Lo Surdo, P, M Iannuccelli, S Contino, L Castagnoli, L Licata, G Cesareni, and L Perfetto. 2023. SIGNOR 3.0, the SIGnaling network open resource 3.0: 2022 update. Nucleic Acids Research 51 (D1): D638–D645. issn: 0305-1048. 10.1093/nar/gkac883.

McEwen, Bruce S. 2007. Physiology and neurobiology of stress and adaptation: central role of the brain. Physiological Reviews 87 (3): 873–904. 10.1152/physrev.00041.2006.

Mikheev, Alexander M, Takeshi Nabekura, Amal Kaddoumi, Theo K Bammler, Rama Govindarajan, Mary F Hebert, and Jashvant D Unadkat. 2008. Gene expression profiling of human term placenta. Reproductive Biology and Endocrinology 6:13. 10.1186/1477-7827-6-13.

Mol, Ben WJ, Claire T Roberts, Shakila Thangaratinam, Laura A Magee, Christianne JM De Groot, and G Justus Hofmeyr. 2016. Pre-eclampsia. The Lancet 387 (10022): 999–1011. 10.1016/S0140-6736(15)00070-7.

Myers, Jessica E, Ruben Tuytten, Gary Thomas, Wim Laroy, Kamiel Kas, Griet Vanpoucke, Carl T Roberts, Louise C Kenny, Nigel AB Simpson, and Philip N Baker. 2013. Proteomic profile of preeclampsia in the first trimester of pregnancy. European Journal of Obstetrics & Gynecology and Reproductive Biology 172:53–59. 10.1016/j.ejogrb.2013.10.015.

Otasek, David, John H Morris, Jorge Bouças, Alexander R Pico, and Barry Demchak. 2019. Cytoscape automation: empowering workflow-based network analysis. Genome Biology 20 (1): 1–15. 10.1186/s13059-019-1758-4.

Palei, Ana CT, Frank T Spradley, Junie P Warrington, Eric M George, and Joey P Granger. 2013. Pathophysiology of hypertension in pre-eclampsia: a lesson in integrative physiology. Acta Physiologica 208 (3): 224–233. 10.1111/apha.12106.

Phipps, Elizabeth A, Ravi Thadhani, Thomas Benzing, and S Ananth Karumanchi. 2019. Pre-eclampsia: pathogenesis, novel diagnostics and therapies. Nature Reviews Nephrology 15 (5): 275–289. 10.1038/s41581-019-0119-6.

Powe, Camille E, Richard J Levine, and S Ananth Karumanchi. 2011. Preeclampsia, a disease of the maternal endothelium: the role of antiangiogenic factors and implications for later cardiovascular disease. Circulation 123 (24): 2856–2869. 10.1161/CIRCULATIONAHA.109.853127.

Ritchie, Matthew E, Belinda Phipson, D. Wu, Yifang Hu, Charity W Law, Wei Shi, and Gordon K Smyth. 2015. Limma powers differential expression analyses for rna-sequencing and microarray studies. Nucleic Acids Research 43 (7): e47. 10.1093/nar/gkv007.

Romero, Roberto, Offer Erez, Maik Huttemann, Eli Maymon, Bogdan Panaitescu, Agustin Conde-Agudelo, et al. 2017. Metformin, the aspirin of the 21st century: its role in gestational diabetes mellitus, prevention of preeclampsia and cancer, and the promotion of longevity. American Journal of Obstetrics and Gynecology 217 (3): 282–302. 10.1016/j.ajog.2017.06.003.

Santillo, M., A. Colantuoni, P. Mondola, B. Guida, and S. Damiano. 2015. Nox signaling in molecular cardiovascular mechanisms involved in the blood pressure homeostasis. Frontiers in Physiology 6:194. 10.3389/fphys.2015.00194.

Shannon, Paul, Andrew Markiel, Owen Ozier, Nitin S Baliga, Jonathan T Wang, Daniel Ramage, Nada Amin, Benno Schwikowski, and Trey Ideker. 2003. Cytoscape: a software environment for integrated models of biomolecular interaction networks. Genome Research 13 (11): 2498–2504. 10.1101/gr.1239303.

Sitras, Vasilis, Ruth H Paulssen, Hege Grønaas, Jørgen Leirvik, Trond A Hanssen, Åse Vårtun, and Ganesh Acharya. 2009. Differential placental gene expression in severe preeclampsia. Placenta 30 (5): 424–433. 10.1016/j.placenta.2009.01.012.

Staff, Anne Cathrine, Stephen J Benton, Peter von Dadelszen, James M Roberts, Robert N Taylor, Rebecca W Powers, D Stephen Charnock-Jones, and Christopher WG Redman. 2013. The two-stage placental model of preeclampsia: an update. Journal of Reproductive Immunology 99 (1-2): 1–15. 10.1016/j.jri.2013.05.003.

Szklarczyk, Damian, Rebecca Kirsch, Maryam Koutrouli, Katerina Nastou, Farrokh Mehryary, Radja Hachilif, Annika L Gable, Tao Fang, Nadezhda T Doncheva, Sampo Pyysalo, et al. 2023. The string database in 2023: protein-protein association networks and functional enrichment analyses for any sequenced genome of interest. Nucleic Acids Research 51 (D1): D638–D646. 10.1093/nar/gkac1000.

Turco, Margherita Y, Lucy Gardner, Richard G Kay, Robert S Hamilton, Marvin Prater, Tereza A Edgell, Amanda M de Mestre, Abigail L Fowden, Ashley Moffett, and Graham J Burton. 2018. Trophoblast organoids as a model for maternal–fetal interactions during human placentation. Nature 564 (7735): 263–267. 10.1038/s41586-018-0753-3.

Vento-Tormo, Roser, Mirjana Efremova, Rachel A Botting, Margherita Y Turco, Miquel Vento-Tormo, Kerstin B Meyer, Jong-Eun Park, Emily Stephenson, Krzysztof Polański, Angela Goncalves, et al. 2018. Single-cell reconstruction of the early maternal–fetal interface in humans. Nature 563 (7731): 347–353.

Wang, Rui-Sheng, Assieh Saadatpour, and Réka Albert. 2012. Boolean modeling in systems biology: an overview of methodology and applications. Physical Biology 9 (5): 055001. 10.1088/1478-3975/9/5/055001.

Weissgerber, Tracey L, and Lanay M Mudd. 2015. Preeclampsia and diabetes: a worrying combination. Current Hypertension Reports 17 (3): 1–10. 10.1007/s11906-015-0543-3.

Xie, Zichen, Avi Bailey, Maxim V Kuleshov, Daniel JB Clarke, John E Evangelista, Sherry L Jenkins, Alexander Lachmann, Marcin L Wojciechowicz, Eliza Kropiwnicki, Kathleen M Jagodnik, et al. 2021. Enrichr: a comprehensive gene set enrichment analysis web server 2021 update. Nucleic Acids Research 49 (W1): W246–W251. 10.1093/nar/gkab302.

Zeng, Heng, Xiaoqing He, Qin-Hui Tuo, DF Liao, Gaol-Qiang Zhang, and Jian-Xiong Chen. 2017. Akt and pathological hypertrophy. International Journal of Cardiology 236:201–212. 10.1016/j.ijcard.2017.01.036.

Zhu, Chenlin, Haitian Yang, Qianzhen Geng, Qiang Ma, Yan Long, Chengwen Zhou, and Ming Chen. 2016. Oxidized low density lipoprotein, maternal hypercholesterolemia and preeclampsia. Lipids in Health and Disease 15 (1): 1–6. 10.1186/s12944-016-0280-x.

